# Validation of a commercially available SARS-CoV-2 serological Immunoassay

**DOI:** 10.1101/2020.05.02.20080879

**Authors:** Benjamin Meyer, Giulia Torriani, Sabine Yerly, Lena Mazza, Adrien Calame, Isabelle Arm-Vernez, Gert Zimmer, Thomas Agoritsas, Jérôme Stirnemann, Hervé Spechbach, Idris Guessous, Silvia Stringhini, Jérôme Pugin, Pascale Roux-Lombard, Lionel Fontao, Claire-Anne Siegrist, Isabella Eckerle, Nicolas Vuilleumier, Laurent Kaiser, for the Geneva Center for Emerging Viral Diseases

## Abstract

**Objectives:** To validate the diagnostic accuracy of a Euroimmun SARS-CoV-2 IgG and IgA immunoassay for COVID-19.

**Methods:** In this unmatched (1:1) case-control validation study, we used sera of 181 laboratory-confirmed SARS-CoV-2 cases and 176 controls collected before SARS-CoV-2 emergence. Diagnostic accuracy of the immunoassay was assessed against a whole spike protein-based recombinant immunofluorescence assay (rIFA) by receiver operating characteristic (ROC) analyses. Discrepant cases between ELISA and rIFA were further tested by pseudo-neutralization assay.

**Results:** COVID-19 patients were more likely to be male and older than controls, and 50.3% were hospitalized. ROC curve analyses indicated that IgG and IgA had high diagnostic accuracies with AUCs of 0.992 (95% Confidence Interval [95%CI]: 0.986-0.996) and 0.977 (95%CI: 0.963-0.990), respectively. IgG assays outperformed IgA assays (p=0.008). Taking an assessed 15% inter-assay imprecision into account, an optimized IgG ratio cut-off > 1.5 displayed a 100% specificity (95%CI: 98–100) and a 100% positive predictive value (95%CI: 97-100). A 0.5 cut-off displayed a 97% sensitivity (95%CI: 93–99) and a 97% negative predictive value (95%CI: 93–99). Substituting these thresholds for the manufacturer’s, improved assay performance, leaving 12% of IgG ratios indeterminate between 0.5-1.5.

**Conclusions:** The Euroimmun assay displays a nearly optimal diagnostic accuracy using IgG against SARS-CoV-2 in patient samples, with no obvious gains from IgA serology. The optimized cut-offs are fit for rule-in and rule-out purposes, allowing determination of whether individuals in our study population have been exposed to SARS-CoV-2 or not. IgG serology should however not be considered as a surrogate of protection at this stage.

## Introduction

High throughput and reliable serological assays detecting antibodies against SARS-CoV-2 are essential to determine the proportion of SARS-CoV-2 infected individuals and estimate the current seroprevalence in the general population or in high-risk groups, such as health care workers. Serological assays can complement diagnostic strategies focusing on the identification of the infectious agent during the acute phase of disease. Unlike RT-PCR, they can identify infected individuals that remained asymptomatic or undiagnosed, which are both frequent conditions during SARS-CoV-2 infection, long after the initial infection. Validated serological assays are also key to understanding the (immuno)-pathophysiology of COVID-19 in various patients’ groups and will be critical to characterize responses elicited by the numerous vaccine candidates in development (1).

Designing serological testing strategies with high sensitivity and specificity and with acceptable positive (PPV) and negative predictive values (NPV) is far from trivial, and requires taking two major analytical aspects into account: analytical specificity and sensitivity (2),(3). The former may largely be determined by the degree of cross-reactivity with other CoVs, which frequently cause common colds in humans (i.e. HCoV-229E, −NL63, −OC43 and −HKU1) (4) resulting in seroprevalence rates usually above 90% in adults (5).

This cross-reactivity occurs when virus-specific antigenic epitopes are highly similar and recognized by the same B cells. It is best defined by the proportion of “false-positive” SARS-CoV-2 results in individuals who were never exposed to this pathogen. In contrast to common cold CoVs, the seroprevalence for MERS-CoV is low even in endemic countries (6). Therefore, cross-reactivity between MERS-CoV and SARS-CoV-2 is not a critical factor when assessing population seroprevalence. Previous studies have shown that antibodies against common cold CoVs can cause considerable cross-reactivity in serological assays, depending on the type of assay and antigens used. Particularly, whole virus- or nucleocapsid protein-based assays showed a higher cross-reactivity compared to whole spike or S1 domain-based assays resulting in lower specificity (4). During the early phase of an outbreak, when SARS-CoV-2 seroprevalence is low, serological testing strategies must have a very high specificity to reach a high positive predictive value (PPV) and avoid false positive results.

On the other hand, analytical sensitivity is strongly influenced by the epidemic course, the disease biology, and numerous analytical factors. All these interrelated items are primarily influenced by the intrinsic immunogenicity of the SARS-CoV-2 antigens and the magnitude and duration of B cell responses elicited by infection, be it asymptomatic, benign, moderate or severe (2),(3).

Last but not least in the context of a pandemic, the availability of high throughput and reliable diagnostic platforms is key for health care systems to effectively handle the testing demand, while respecting clinically compatible diagnostic turnaround times (TAT). In this study, we performed an extensive validation of a high throughput SARS-CoV-2 commercial serological platform quantifying both serum IgG and IgA against the S1 protein. As reference method, we used a whole spike-based recombinant immunofluorescence assay (rIFA) (4),(7),(8). Selected sera from SARS-CoV-2-infected patients were assessed for their neutralization capacity using a pseudovirion assay (see below).

## Methods

### Aims

The aim of this study is to validate and define the operational cut-off values of a commercially available ELISA-based SARS-CoV-2 serological assay that could be applied at large scales to reliably determine the presence of specific IgG as a marker of SARS-CoV-2 infection. This study used RT-PCR confirmed cases, but the goal was to be able to identify exposure to SARS-CoV-2 by immunoassay alone. Therefore, the ELISA results were compared against recombinant immunofluorescence assay (rIFA), which was considered as the reference method due to its demonstrated high specificity for serology of other CoVs such as MERS-CoV (6). The secondary goal is to assess the potential added value of IgA in patients recently infected with SARS-CoV-2.

### Study population

Negative control serum samples (n=176) were collected for various serological testing in our routine laboratory and stored for analytical validation. These sera were collected in 2013, 2014 and 2018 before the start of the outbreak and thus have not been exposed to SARS-CoV-2. Sera (n=181) of PCR-confirmed COVID-19 patients were collected at the University Hospitals of Geneva from hospitalized patients (n=91) as well as from patients from the outpatient clinic (n=90). Ethical approval for all sera used in this study was waived by the local ethics committee of the HUG that approves usage of leftover of patient serum collected for diagnostic purposes. The number of days from symptom onset to blood collection was based on patient history whenever this information was available or could be retrieved in a reliable way; otherwise, we used the date of RT-PCR positivity as a surrogate for onset of symptoms. Serum samples from unmatched PCR-confirmed COVID-19 hospitalized patients were collected for routine diagnostic purposes under a general informed consent and outpatients were asked if they were willing to return to the hospital after their symptoms had subsided and to donate a serum sample under written general informed consent.

### Blood sample collection

We used serum samples collected on patient admission, during hospitalization or, when needed, left over sera used for other routine investigations. Samples were immediately processed and then frozen and stored at −20°C until ELISA, recombinant immunofluorescence analyses, and pseudoneutralization were performed (see below). Patients were sampled at different times after onset of symptoms (dpos) or according to days post RT-PCR diagnosis (dpd) if the onset was not known. To compare the seropositivity between different time points, we grouped patients in the following dpos/dpd categories: 0–10 (n=8, 4.4%), 11–20 (n=112, 61.9%) and 21–39 (n=61, 33.7%).

### SARS-CoV-2 RT-PCR analyses

SARS-CoV-2 RT-PCR was performed according to manufacturers’ instructions on various platforms, including initially in house method using eMAG (bioMérieux, France) and Charité RT-PCR protocol(9), then BD SARS-CoV-2 reagent kit for BD Max system (Becton, Dickinson and Co, US) and Cobas 6800 SARS CoV2 RT-PCR (Roche, Switzerland).

### Serum IgG and IgA ELISA

Both IgG and IgA enzyme-linked immunosorbent assays (ELISA) used the S1-domain of the spike protein of SARS-CoV-2 as antigen. Sera were diluted at 1:101 and assessed with the IgG and IgA CE-marked ELISA (Euroimmun AG, Lübeck, Germany # EI 2606-9601 G and # EI 2606–9601 A) according to the manufacturer’s instructions. They were run on Dynex Agility (Ruwag, Switzerland) according to the manufacturer’s protocol. After adding the conjugate, samples’ immunoreactivity was measured at an optical density of 450nm (0D450) and then divided by the 0D450 of the calibrator provided with each ELISA kit to minimize inter-assay variation(8). The quantitative results obtained were then expressed in arbitrary units and interpreted as follows: OD ratio: <0.8 = negative; ≥0.8 and <1.1 = indeterminate; ≥1.1 = positive. Inter-assay variation was 15.6% for IgG at a ratio of 2.09 (n=17) and 17.7% for IgA at a ratio of 4.85 (n=7).

### Recombinant immunofluorescence assay

The IgG antibody response against the spike protein of SARS-CoV-2 was assessed by rIFA as described and previously validated for MERS-CoV (6),(7). Briefly, Vero B4 cells were transfected with the mammalian expression vector pCG1-SCoV2-S (kindly provided by M. Hoffmann and S. Pöhlmann, DPZ, Göttingen, Germany) using Fugene HD (Promega, #E2311). After 24h of incubation cells were detached and residual trypsin was removed by centrifugation at 300x g for 5min. 50μl of transfected cells were seeded at a density of 2×10^5^/ml on multi-test glass slides (DUNN Labortechnik GmbH #40-412-05) and incubated for 6h at 37°C, 5% CO_2_. Afterwards slides were washed 2x with PBS and fixed for 10min using ice-cold Acetone/Methanol (ratio 1:1). For rIFA staining, slides were rehydrated for 10min using PBS + 0.1% Tween20 (PBS-T) and blocked with 5% milk in PBS-T for 30min at RT. Sera were diluted 1:40 in blocking buffer, 30μl were applied on each spot and incubated for 60min at 37°C or RT. After 3x washing with PBS-T, secondary goat anti-human-IgG antibody conjugated with Alexa488 (Jackson ImmunoResearch, #109-545-088) was diluted 1:200 in PBS and 25μl were applied to each spot. The secondary antibody was incubated for 45min at 37°C and slides were washed 3x with PBS-T afterwards. Slides were briefly rinsed with dH_2_O and mounted using glycerol. rIFA results were judged by 3 observers independently and the inter-observer kappa correlation was 0.59 (95%CI: 0.31-0.87) between observer 1 and 2, 0.62 (95%CI: 0.33-0.91) between observer 2 and 3 as well as 0.75 (95%CI: 0.52-0.99) between observer 1 and 3, which could be considered a substantial level of agreement(10).

### VSV-based pseudo-neutralization assay

VeroE6 cells were seeded in 96-well plates at 2 × 10^4^ cells per well and grown into confluent monolayer overnight. Sera from patients were inactivated at 56°C for 30 minutes and diluted 1:5 in 120 μl DMEM 2% FCS in the first column of a 96-well plates (in duplicate). Remaining wells were filled with 60 μl of DMEM 2% FCS. Two-fold dilutions were performed until 1:80 was reached by moving 60 μl from one well column to the following one. VSV-based SARS-CoV-2 pseudotypes (generated according to Berger, Rentsch, and Zimmer (11) and Torriani et al. (12)) expressing a 19 amino acids C-terminal truncated spike protein (13) (NCBI Reference sequence: NC_045512.2) were diluted in DMEM 2% FCS in order to have MOI=0.01 in 60 μl volume and added on top of serum dilutions (final serum dilutions obtained were from 1:10 to 1:160). The virus-serum containing plate was incubated at 37°C, for 2h. Vero E6 were then infected with 100μl of virus-serum mixtures. After incubation at 37°C for 1.5h, cells were washed once with 1X PBS and DMEM 10% FCS was added. After 16-20h of incubation at 37°C, 5% CO2 cells were fixed with 4% formaldehyde solution for 15 min at 37°C and nuclei stained with 1μg/ml DAPI (AppliChem #A4099) solution. GFP positive infected cells were counted with ImageXpress® Micro Widefield High Content Screening System (Molecular Devices) and data analyzed with MetaXpress 5.1.0.41 software.

### Statistical analyses

Analyses were performed with Graph Pad Prism version 8.3.1 software using Fisher’s bilateral exact test and Mann-Whitney *U*-test where appropriate. ROC analyses were performed using analyse-it™ software for Excel (Microsoft, Redmond, WA, USA) to: (i) confirm the cut-off values, prospectively proposed by the manufacturer, when compared to IFA results as the reference method (samples with very weak fluorescence signal just above background were deemed putative negatives, and were considered as negative for the purpose of this analysis); (ii) compare the diagnostic accuracy of IgG alone, IgA alone, or both combined; and (iii) determine the optimal rule-in cut-off (PPV of 100%), a rule-out cut-off with a NPV above 95%, and an indeterminate interval, taking into account the analytical imprecision-derived least significant change (LSC). The LSC represents the smallest significant detectable difference between two measurements based given the analytical imprecision, and is conventionally defined as 1.96 * √2 * VC,(14),(15). AUC comparisons were performed according to the nonparametric approach proposed by DeLong *et al*. (16). Sensitivity (SE), specificity (SP), PPV and NPV with the respective 95% CIs are given. A value of p<0.05 was considered statistically significant.

## Results

The baseline demographic characteristics of the participants are summarized in table 1. There was a higher proportion of males (61.3%) vs females (38.7%) in COVID-19 patients compared to controls (Fisher’s exact test, p=0.002). Among COVID-19 patients, we included an equal representation of hospitalized patients (n=91, 50.3%) and outpatients (n=90, 49.7%).

**Table 1.**

|  | Negative Controls |  |  | COVID-19 Patients |  |  |
| --- | --- | --- | --- | --- | --- | --- |
|  | No (%) | Median (IQR) | Range | No (%) | Median (IQR) | Range |
| Age (years) |  |  |  |  |  |  |
| All | 176 (100) | 39.2 (23.8) | 18.0-89.5 | 181 (100) | 53.1 (29.2) | 20.1-86.9 |
| 18-40 | 100 (56.8) |  |  | 61 (33.7) |  |  |
| 41-65 | 61 (34.7) |  |  | 70 (38.7) |  |  |
| 66+ | 15 (8.5) |  |  | 50 (27.6) |  |  |
| Gender |  |  |  |  |  |  |
| Female | 97 (55.1) |  |  | 70 (38.7) |  |  |
| Male | 79 (44.9) |  |  | 111 (61.3) |  |  |
| Hospitalized |  |  |  | 91 (50.3) |  |  |
| Outpatient |  |  |  | 90 (49.7) |  |  |
| DPOS/DPD |  |  |  |  |  |  |
| 0-10 |  |  |  | 8 (4.4) | 7.0 (1.75) | 0-8 |
| 11-20 |  |  |  | 112 (61.9) | 15.0 (2.0) | 11-20 |
| 21-39 |  |  |  | 61 (33.7) | 28.0 (5.0) | 21-39 |

DPOS days post onset of symptoms, DPD days post diagnosis, IQR inter quartile range

### Prevalence of IgG seropositivity according to rIFA

An in-house developed recombinant immunofluorescence assay (rIFA) using the whole spike protein of SARS-CoV-2 as antigen was used to assess spike-specific serum IgG. Since rIFA was used as a confirmatory assay, we maximized its specificity by interpreting putative negatives as negative to avoid false positive results. Among negative control samples, we found no positive, five putative negative (2.8%) and 171 (97.2%) negative samples indicating a specificity of 100%. Among COVID-19 samples there were 165 (91.2%) positive, two (1.1%) putative negative and 14 (7.7%) negative samples, indicating an overall detection rate of 91.2%. We found that rIFA seropositivity was low at 0-10 dpos/dpd (12.5%) but increased to 92.0% and 100% in sera collected at 11-20 and 21-39 dpos/dpd, respectively (table 2). Similar results were found in hospitalized and outpatients. Thus, in our sample of 181 COVID-19 samples, rIFA proved a robust method for the detection of SARS-CoV-2 spike-specific IgG.

**Table 2.**

| Samples | Total No. (%) | S-rIFA IgG No. (%) |  |  | ELISA IgG No. (%) EI cut-offs<sup>1</sup> |  |  | ELISA IgG No. (%) GE cut-offs<sup>2</sup> |  |  |
| --- | --- | --- | --- | --- | --- | --- | --- | --- | --- | --- |
|  |  | Negative | Putative Neg. | Positive | Negative | Indeterminate | Positive | Negative | Indeterminate | Positive |
| Negative Controls | 176 (100) | 171 (97.2) | 5 (2.8) | 0 (0) | 171 (97.2) | 5 (2.8) | 0 (0) | 156 | 20 (11.4) | 0 (0) |
| COVID-19 Patient |  |  |  |  |  |  |  |  |  |  |
| All | 181 (100) | 14 (7.7) | 2 (1.1) | 165 (91.2) | 26 (14.4) | 1 (0.6) | 154 (85.1) | 15 (8.3) | 23 (12.7) | 143 (79.0) |
| 0-10 DPOS / DPD | 8 (4.4) | 6 (75.0) | 1 (12.5) | 1 (12.5) | 8 (100) | 0 (0) | 0 (0) | 7 (87.5) | 1 (12.5) | 0 (0) |
| 11-20 DPOS / DPD | 112 (61.9) | 8 (7.1) | 1 (0.9) | 103 (92.0) | 17 (15.2) | 0 (0) | 95 (84.8) | 7 (6.3) | 19 (17.0) | 86 (76.8) |
| 21-39 DPOS / DPD | 61 (33.7) | 0 (0) | 0 (0) | 61 (100) | 1 (1.6) | 1 (1.6) | 59 (96.7) | 1 (1.6) | 3 (4.9) | 57 (93.4) |
| COVID-19 Outpatients |  |  |  |  |  |  |  |  |  |  |
| All | 90 (100) | 8 (8.9) | 0 (0) | 82 (91.1) | 14 (15.6) | 0 (0) | 76 (84.4) | 7 (7.8) | 14 (15.6) | 69 (76.6) |
| 0-10 DPOS / DPD | 5 (5.6) | 4 (80) | 0 (0) | 1 (20) | 5 (100) | 0 (0) | 0 (0) | 4 (80.0) | 1 (20.0) | 0 (0) |
| 11-20 DPOS / DPD | 41 (45.6) | 4 (9.8) | 0 (0) | 37 (90.2) | 8 (19.5) | 0 (0) | 33 (80.5) | 2 (4.9) | 11 (26.8) | 28 (68.3) |
| 21-39 DPOS / DPD | 44 (48.8) | 0 (0) | 0 (0) | 44 (100) | 1 (2.3) | 0 (0) | 43 (97.7) | 1 (2.3) | 2 (4.6) | 41 (93.2) |
| COVID-19 Hospitalized |  |  |  |  |  |  |  |  |  |  |
| All | 91 (100) | 6 (6.6) | 2 (2.2) | 83 (91.2) | 12 (13.2) | 1 (1.1) | 78 (85.7) | 8 (8.8) | 9 (9.9) | 74 (81.3) |
| 0-10 DPOS / DPD | 3 (3.3) | 2 (66.7) | 1 (33.3) | 0 (0) | 3 (100) | 0 (0) | 0 (0) | 3 (100) | 0 (0) | 0 (0) |
| 11-20 DPOS / DPD | 71 (78.0) | 4 (5.6) | 1 (1.4) | 66 (93.0) | 9 (12.7) | 0 (0) | 62 (87.3) | 5 (7.0) | 8 (11.3) | 58 (81.7) |
| 21-39 DPOS / DPD | 17 (18.7) | 0 (0) | 0 (0) | 17 (100) | 0 (0) | 1 (5.9) | 16 (94.1) | 0 (0) | 1 (5.9) | 16 (94.1) |

DPOS days post onset of symptoms, DPD days post diagnosis, rIFA recombinant immunofluorescence assay
1: EI cut-offs: <0.8 = negative; ≥0.8 and <1.1 = indeterminate; ≥1.1 = positive
2: GE cut-offs: <0.5 = negative; ≥0.5 and <1.5 = indeterminate; ≥1.5 = positive

### Diagnostic accuracies of IgG and IgA ELISA against rIFA

Overviews of OD ratios for IgG and IgA are shown in Figure 1 A and B, respectively. In control sera, we found no positive, five (2.8%) indeterminate and 171 (97.2%) negative samples for IgG, and 14 (7.7%) positive, 10 (5.7%) indeterminate and 157 (86.7%) negative samples for IgA according to the manufacturer defined cut-off (EI cut-offs) (table 2). Considering the indeterminate values as positive resulted in a specificity of 97.3% (IgG) and 86.4% (IgA), respectively. Analysis of COVID-19 samples revealed 154 (85.1%) positive, one (0.6%) indeterminate and 26 (14.4%) negative samples for IgG, and 164 (90.6%) positive, four (2.2%) indeterminate and 13 (7.2%) negative samples for IgA. This resulted into an overall significantly higher seropositivity for IgA (90.6%) than IgG (85.1%) in the S1-based ELISA (p= 0.041).

**Figure 1:**
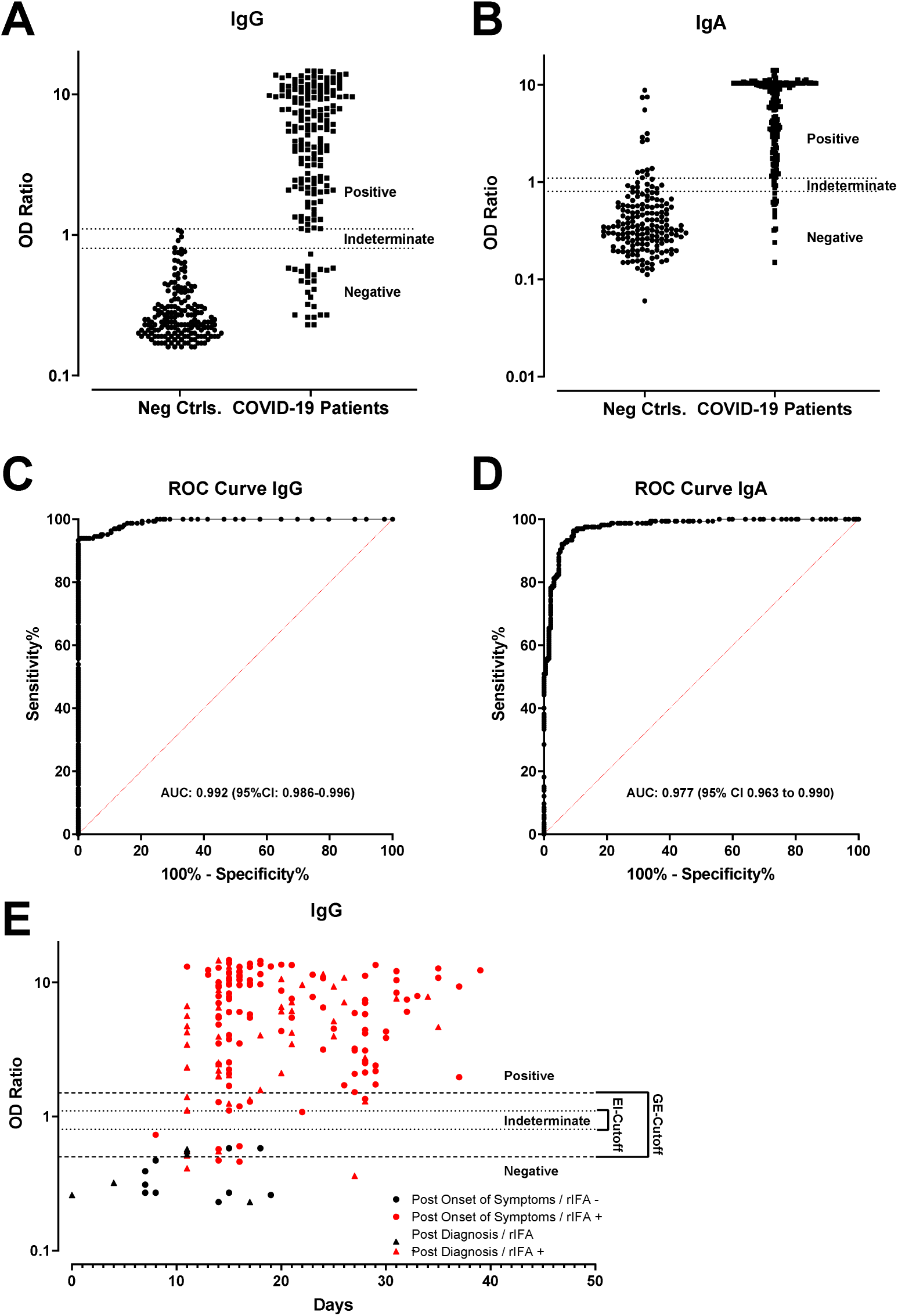
**A** and **B**: OD ratio of negative control samples (n=176) and PCR-confirmed CoVID-19 patients (n=181) were determined using an IgG and IgA ELISA. C and D: ROC curve analysis for IgG and IgA OD ratio results including the area under the curve (AUC) and 95% confidence intervals. E IgG OD ratios of COVID-19 patients at different days post onset of symptoms (dpos, dots) or days post diagnosis (dpd, triangles). Red dots or triangles show samples that are confirmed by whole spike recombinant immunofluorescence analysis. Dotted and dashed lines indicate the Euroimmun-(EI) or Geneva-(GE) cut-offs for negative, indeterminate and positive samples, respectively.

Next, we analysed the seropositive rates of both ELISAs at different dpos or dpd (Figure 1E) according to the EI cut-offs. Sera collected 21 dpos/dpd had a similarly high seropositivity for both IgG (96.7%) and IgA (96.7%) (p >0.99). A higher seropositivity was observed for IgA compared to IgG for sera collected at 11-20 dpos/dpd (91.1% vs 84.8%, p= 0.0264) but not 0-10 dpos/dpd (37.5% vs 0%, p= 0.2). No significant difference was found between hospitalized and outpatients in IgG or IgA ELISA (p= 0.833, and p= 1.000, respectively).

ROC curve analysis (Fig 1; C and D) indicated that overall, both IgG and IgA had a high diagnostic accuracy with respective AUCs of 0.992 (95%CI: 0.986-0.996) and 0.977 (95%CI: 0.963-0.99), respectively. Although modest, this AUC difference was found to be significant according to the de Delong method (p=0.008). Combining IgG and IgA ELISAs together (AUC 0.986; 95%CI: 0.976-0.995) tended to decrease the diagnostic accuracy of the model when compared to IgG alone (p=0.080), but outperformed IgA alone (p<0.001). Although these differences were modest, our results indicate that IgG ELISA displays the optimal fit with IgG rIFA and that IgA did not improve diagnostic value. In a subgroup analysis considering only sera harvested before 21 days after symptoms onset (n=120), ROC curve analyses displayed similar results - with an AUC of 0.981 for the IgG ratio and of 0.965 for the IgA ratio. At these early time-points, the AUC difference between IgG and IgA ratio was not significant (p=0.19).

Thirteen patient samples had discrepant or potentially discrepant results, i.e. were negative or indeterminate in ELISA and putative negative or positive in rIFA. To investigate this discordance, we performed a VSV-based pseudovirus neutralization assay (pseudo-NT) for 1) these 13 samples (n=13), 2) five negative control samples, including one sample that showed putative negative results in rIFA, and 3) seven patient samples that were positive by ELISA IgG, IgA and rIFA (table 3). All samples that showed negative and putative negative rIFA results were confirmed negative, whereas all rIFA positive samples were confirmed as positive by the pseudo-NT assay (table 3). These findings support the accuracy of the rIFA and our decision to count putative negative rIFA results as negative.

**Table 3.**

| Sample Code | DPOS/DPD | Type | Result |  |  | Reciprocal Endpoint Titre |  |
| --- | --- | --- | --- | --- | --- | --- | --- |
|  |  |  | ELISA IgG | S-rIFA IgG | ELISA IgA | Pseudo-NT 90 | Pseudo-NT 50 |
| 8 | NA | Neg. Ctrl. | 0.79 | NEG | 1.29 | <10 | <10 |
| 9 | NA | Neg. Ctrl. | 0.31 | NEG | 0.56 | <10 | <10 |
| 10 | NA | Neg. Ctrl. | 0.57 | NEG | 0.79 | <10 | <10 |
| 11 | NA | Neg. Ctrl. | 0.97 | NEG | 2.72 | <10 | <10 |
| 56 | NA | Neg. Ctrl. | 0.81 | Putative NEG | 0.47 | <10 | <10 |
| 209 | 8 | COVID-19 Pat. | 0.73 | POS | 3.66 | 20 | ≥160 |
| 30181704 | 22 | COVID-19 Pat. | 1.08 | POS | 0.77 | 20 | ≥160 |
| 30189617 | 16 | COVID-19 Pat. | 0.60 | POS | 9.99 | 20 | 80 |
| 30186069 | 14 | COVID-19 Pat. | 0.55 | POS | 2.94 | 20 | 80 |
| 30193397 | 11 | COVID-19 Pat. | 0.56 | POS | 4.21 | 10 | 80 |
| 30193613 | 11 | COVID-19 Pat. | 0.51 | POS | 0.92 | 80 | ≥160 |
| 30193717 | 11 | COVID-19 Pat. | 0.41 | POS | 1.02 | 10 | 40 |
| 30197451 | 14 | COVID-19 Pat. | 0.57 | POS | 3.85 | ≥160 | ≥160 |
| 30197527 | 16 | COVID-19 Pat. | 0.46 | POS | 0.67 | <10 | 20 |
| 30202986 | 18 | COVID-19 Pat. | 0.58 | Putative NEG | 1.53 | <10 | <10 |
| 30208864 | 14 | COVID-19 Pat. | 0.47 | POS | 0.33 | 20 | 80 |
| 30208885 | 7 | COVID-19 Pat. | 0.39 | Putative NEG | 0.48 | <10 | <10 |
| 30251875 | 27 | COVID-19 Pat. | 0.36 | POS | 0.51 | 10 | 80 |
| 202 | 16 | COVID-19 Pat. | 6.02 | POS | 10.70 | 160 | ≥160 |
| 205 | 11 | COVID-19 Pat. | 13.08 | POS | 10.93 | ≥160 | ≥160 |
| 30175134 | 15 | COVID-19 Pat. | 14.64 | POS | 10.41 | ≥160 | ≥160 |
| 30175147 | 15 | COVID-19 Pat. | 13.90 | POS | 10.41 | ≥160 | ≥160 |
| 30175152 | 18 | COVID-19 Pat. | 13.67 | POS | 10.41 | ≥160 | ≥160 |
| 30175157 | 15 | COVID-19 Pat. | 9.74 | POS | 9.65 | ≥160 | ≥160 |
| 30193442 | 11 | COVID-19 Pat. | 1.40 | POS | 6.48 | 20 | 80 |

DPOS days post onset of symptoms, DPD days post diagnosis, rIFA recombinant immunofluorescence assay, NA not applicable, Pseudo-NT 90 and 50: pseudovirus neutralization test using 90% or 50% reduction as cut-off

### Optimizing the threshold for IgG serology

Taking into account the variation coefficient (VC) of 15% at a low positivity IgG ratio (2.09), and assuming that such VC could at least be transposed to the 1.1 cut-off ratio, the minimal LSC would be 0.42. Added to the 1.1 ratio, an IgG ratio cut-off of 1.5 (1.1 + 0.42) would be the lowest able to secure 100% PPV while taking into account the analytical imprecision. Furthermore, an LSC of 0.42 also indicates that the indeterminate zone proposed by the manufacturer (between 0.8 and 1.1) should be reconsidered, as it falls within the analytical imprecision range. At the 1.5 IgG ratio cut-off, ROC curve analyses indicated that the SE was 86%, the SP 100%, and the NPV 89%. Thus, selecting a 1.5 cut-off, rather than the recommended 1.1 cut-off for IgG seropositivity, allows the securing of a PPV of 100% despite a 15% imprecision. Higher VCs would translate into a higher seropositivity cut-off to secure an identical PPV. For rule-out purposes (i.e. the seronegativity lower cut-off), the best trade-off IgG ratio cut-off was found to be < 0.5. At this value, the SE was 97%, the SP 87%, the NPV 97% and the PPV 86% (table 4). This defines an indeterminate range between IgG ratios of 0.5 and 1.5, which represented 43 cases (12%) of our samples (including 20 control and 23 COVID-19 samples). In this indeterminate zone, all 20 sera from controls were confirmed as negative (n=19) or putative negative (n=1) by rIFA. For indeterminate samples of COVID-19 patients, five were negative, one was a putative negative and 18 were positive by rIFA.

**Table 4.**

|  | Cut-Offs | Sensitivity (95%) | Specificity (95%) | Positive predictive value (95%) | Negative predictive value (95%) |
| --- | --- | --- | --- | --- | --- |
| **All samples (n=357)**<br>**IgG ratio**<br>**AUC: 0.992**<br>**(95%CI: 0.986-0.996)** | GE cut-offs<sup>1</sup> |  |  |  |  |
|  | >1.5 (rule-in) | 86% (80-91) | 100% (98-100) | 100% (97-100) | 89% (84-93) |
|  | <0.5 (rule-out) | 97% (93-99) | 87% (81-91) | 86% (81-90) | 97% (93-99) |
|  | EI cut-offs<sup>2</sup> |  |  |  |  |
|  | >1.1 (rule in) | 93% (87-96) | 100% (98-100) | 100% (97-100) | 94% (90-97) |
|  | <0.8 (rule out) | 94% (88-94) | 98% (94-99) | 97% (92-99) | 95% (91-97) |
| **<21DPOS/DPD(n=120)**<br>**IgG ratio**<br>**AUC: 0.981**<br>**(95%CI: 0.961-1.00)** | GE cut-offs<sup>1</sup> |  |  |  |  |
|  | >1.5 (rule-in) | 82% (73-88) | 100% (76-100) | 100% (95-100) | 46% (29-63) |
|  | <0.5 (rule-out) | 96% (90-99) | 69% (41-88) | 95% (89-98) | 73% (45-91) |
|  | EI cut-offs<sup>2</sup> |  |  |  |  |
|  | >1.1 (rule in) | 91% (83-95) | 100% (76-100) | 100% (95-100) | 62% (41-79) |
|  | <0.8 (rule out) | 91% (84-96) | 100% (76-100) | 100% (95-100) | 64% (43-81) |

DPOS days post onset of symptoms, DPD days post diagnosis, AUC area under the curve
1: GE cut-offs: <0.5 = negative; ≥0.5 and <1.5 = indeterminate; ≥1.5 = positive
2: EI cut-offs: <0.8 = negative; ≥0.8 and <1.1 = indeterminate; ≥1.1 = positive

In the subgroup of patients whose samples were taken before 21 dpos, using the manufacturer IgG ratio seropositivity cut-off (1.1), the SE was 91%, the SP 100%, the PPV 100%, and the NPV 62%. At the manufacturer seronegative cut-off (<0.8), the SE was 91%, the SP 100%, the PPV 100%, and the NPV 64% (table 4). Importantly, no patient displayed a ratio between 0.8 and 1.1 in this subgroup, preventing us from estimating the importance of the indeterminate cases.

Using the aforementioned optimized IgG ratio cut-off for IgG seropositivity (1.5), the SE was 82%, the SP 100%, the PPV 100%, and the NPV 46%. At the cut-off for IgG seronegativity (<0.5), the SE was 96%, the SP 69%, the PPV 95%, and the NPV 73% (table 4). The number of undetermined cases between 0.5 and 1.5 IgG ratio values represented 17% (20/120) of the subgroup. Taken together, these results indicate that if the seropositivity cut-off of 1.5 for the IgG ratio would be suitable for patients with sample taken <21 days post symptoms onset, the seronegative cut-off of 0.5 would display a non-optimal NPV for rule-out purposes. The optimal seronegative (NPV 100%) cut-off would have been <0.4, at the cost of a modest increase of indeterminate cases to 20% (24/120). Thus, this assay performs best for convalescent samples taken as of 21 dpos.

## Discussion

The key finding of this validation study, derived from a large cohort of laboratory-confirmed SARS-CoV-2 cases and unmatched negative controls, is that the present CE-IVD marked immunoassay for IgG has a good diagnostic accuracy with an AUC of 0.99 and outperformed IgA according to ROC curve comparisons. These results parallel the ones obtained by two other smaller studies (8),(17), which tested the non CE-marked ELISA version and showed potentially better diagnostic performances of IgG compared to IgA ELISA but lower AUCs compared to this study. Our improved results might result from the higher number of negative controls and COVID-19 patient sera tested, and from the decreased non-specific background relative to that observed in the earlier studies. Despite the greater performance of the IgG based ELISA, it was reassuring to see that the IgA results correlated well with the IgG results and gave very similar AUCs upon ROC analyses. However, knowing if and how IgA could be used in clinical practice to further refine rule-in our rule-out strategies still remains elusive.

The second notable finding of this study is that the current manufacturer cut-offs are prone to misinterpretations and should not be used without proper evaluation before routine testing. Keeping in mind the challenges of developing a serological assay for a new viral disease in an emergency situation; securing both rule-in and rule-out cut-offs is key to mitigate the unavoidable risk of false positive and false negative results due to the combination of a highly dynamic pre-test probability variation, with a suboptimal seroconversion time when studies are undertaken at the peak of an epidemic. Our analyses revealed the following limitations of the manufacturer’s seropositivity cut-off. First, with an inter-assay imprecision of 15% assessed at an IgG ratio of 2.09 (above than the 1.5 two cut-off value selected) translating into a LSC of 0.42 IgG ratio, our results indicate that the analytical imprecision is higher than the range of the indeterminate zone proposed by the manufacturer, which encompasses a delta of 0.3 IgG ratio. This implies that any result within the 0.8-1.1 IgG ratio range could be randomly either above, within or below these values just because of analytical imprecision. Secondly, with a LSC of 0.42 our results indicated that a higher IgG ratio cut-off value was needed to secure an optimal specificity and PPV. Adding the 0.42 LSC to the 1.1 cut-off yielded a 1.5 ratio as the IgG seropositivity cut-off with a PPV of 100%, the lower end of the 95%CI still being compatible with a 97% rule-in strategy. Notably, using this cut-off in the subgroup of patients under 21 dpos/dpd, the SP and PPV were still 100%, however, with broader confidence intervals. Similarly, in an attempt to maximize the negative predictive value at the rule-out cut-off, the cut-off had to be decreased from 0.8 to 0.5 IgG ratio in order to reach an overall NPV of 97%, with a 93% at the lower end of the 95CI. In the subgroup of patients under 21 dpos/dpd this interval was found to be substantially larger (95%CI: 45–91). Taken together, these results indicate that at this stage the optimal rule-in cut-off should be set at >1.5 of IgG ratio for seropositivity and at <0.5 of IgG ratio for rule-out purposes (seronegativity). With these optimized “Geneva’s” cut-offs, the indeterminate zone represented 12% of the cases overall (20 controls and 23 COVID-19 samples), representing a volume easily amenable to further confirmation tests like rIFA.

In this respect, the 13 discordant cases between ELISA and rIFA were tested using the pseudo-NT assay. This indicated that all non-positive samples (negative and putative negative) by rIFA were negative in the pseudo-NT assay, and that all positive samples by rIFA (but ELISA negative) were positive in the pseudo-NT assay. One sample that was indeterminate in ELISA and positive in rIFA was also positive in pseudo-NT. These findings may indicate an earlier IgG antibody response, which could better mediate virus neutralization, against the S2 domain of the spike protein, as previously suggested for SARS-CoV-1 neutralizing antibodies targeting the S2 domain (18),(19),(20),(21). Although based upon a limited number of observations which prevents us from drawing final conclusions, these results suggest that rIFA may be an appropriate confirmatory assay in the future.

Regarding potential limitations, we need to highlight several additional points. We evaluated an ELISA assay measuring antibodies against the S1 domain of the spike protein and not against the full protein, which may contain other highly relevant epitopes from the S2 domain. Such factors could potentially explain why some samples were negative by ELISAs but positive by rIFA, as the whole-spike protein is used in rIFA. Secondly, we strongly emphasize two important cut-off limitations related to the analytical imprecision. At the level of the analytical imprecision, the SARS-CoV-2 IgG intra-individual biological variation being unknown, we had to use the LSC instead of the reference change value (22), which most likely would have translated into a higher seropositivity cut-off. Along the same line, because the intra-lot imprecision of the reagents is still undetermined, but expected to be higher than 15%, this could have a similar impact on cut-off determination. Nevertheless, these cut-offs implemented in routine testing at the Geneva University Hospitals (GE-cut-offs) proved useful in the management of a number of clinically compatible COVID-19 patients with negative PCR results.

Importantly, this study is a diagnostic accuracy validation study and not a seroprevalence study. This implies that the current seropositivity cut-off has to be considered with caution in population studies. Indeed, the 100% PPV achieved in this study was due to combined effect of a 100% specificity and a 1:1 distribution of control and samples form patients with positive SARS-CoV-2 PCR. Therefore, in population seroprevalence studies with a lower expected proportion of COVID-19, the PPV at the 1.5 cut-off will likely decrease. In this context, increasing the rule-in IgG cut-off to a higher value or using a secondary specific confirmatory assay may become necessary. Due to this limitation, at this stage we recommend the confirmation of all positive ELISA results (including ratios above 1.5) using a second serological assay such as rIFA for seroprevalence studies with a low pre-test probability, and the confirmation of doubtful ELISA results in settings with a pre-test probability around 50%. A summary of our proposed testing strategy is given in figure 2.

**Figure 2:**
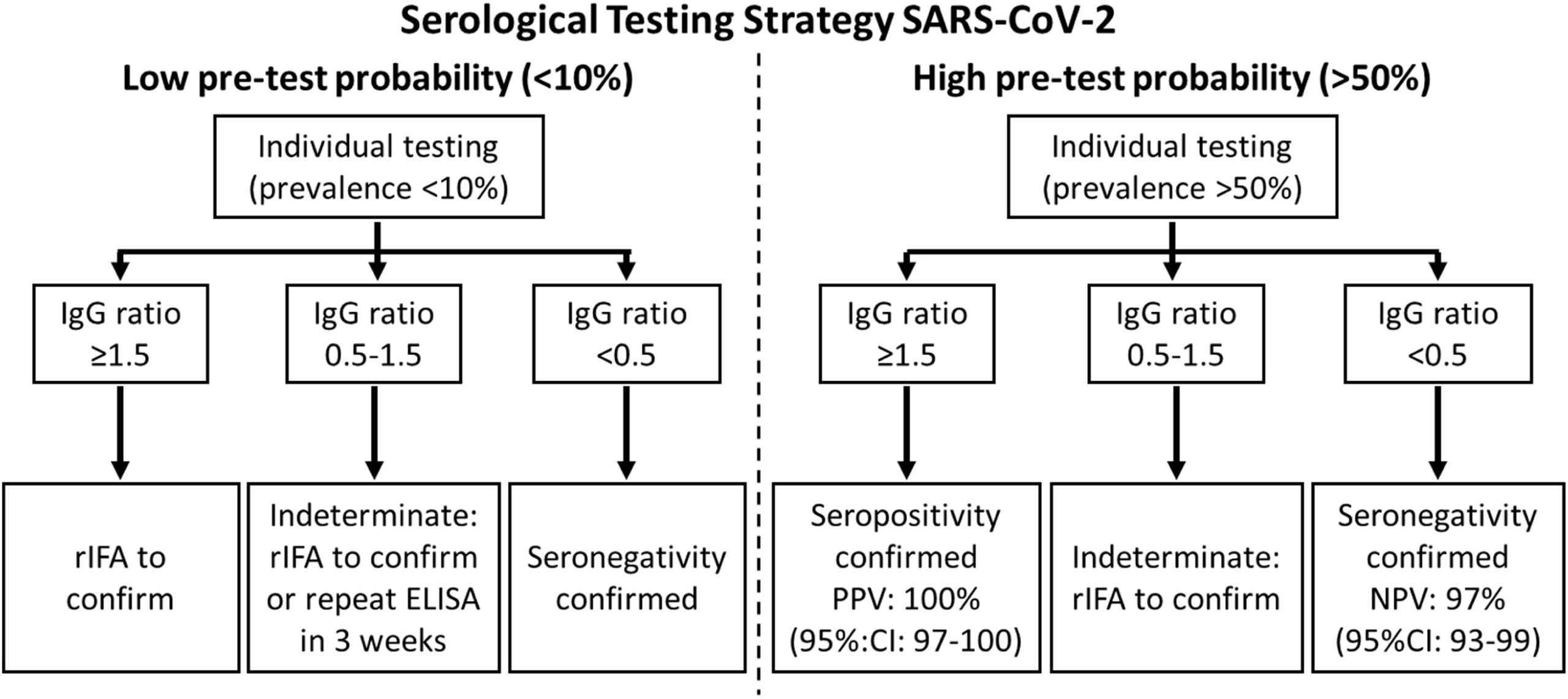
Graphical overview or serological testing strategies in low (<10%) and high (>50%) pre-test probability settings. PPV: positive predictive value; NPV: negative predictive value; rIFA: recombinant immunofluorescence assay; 95%CI: 95% confidence intervals.

In conclusion, in this validation study performed on 357 sera, of which 50.7% came from patients with COVID-19, we demonstrate a close to optimal diagnostic accuracy of IgG SARS-CoV-2 serology of the Euroimmun assay, without any obvious gains from IgA serology. Taking analytical imprecision into account, we propose optimized cut-offs allowing a PPV of 100% and a NPV of 97% to be secured with an indeterminate zone comprising about 12% of the results, for which additional rIFA analyses are currently necessary. Ongoing seroprevalence studies will be instrumental to further refine the optimal rule-in and rule-out cut-offs, as well as the optimal testing strategy, which may require a highly specific confirmatory assay. For the time being, this assay seems to be fit for the purpose of enabling authorities to make informed decisions regarding measures to be taken in order to manage the SARS-CoV-2 pandemic. Using the GE cut-offs could also allow the determination of whether individuals have been exposed or not to SARS-CoV-2 with high confidence. As a final caution, at this time simple IgG levels determined by ELISA or rIFA cannot be considered as a surrogate of protection for individual patients. This implies that risk mitigation decisions may not yet be based only on SARS-CoV-2 ELISA/rIFA serology.

## Data Availability

Data will be made available upon reasonable request or after accepted publication of the manuscript.

## Acknowledgements

We would like to thank Markus Hoffmann and Stefan Pöhlmann (DPZ, University of Göttingen) for providing a SARS-CoV-2 spike protein expression vector. We thank Erik Boehm for help with writing and editing the manuscript. We are also thankful to Romain Burquier, Krystel Tommasini, Karine Derrac, Lydia Massa-Rouffineau, Myriam Florey-Thévenoz, Katia, Marjan Mohitzadeh and Liliane Marchand for technical assistance and help with the serological assays. Last, we would like to thank patients for providing samples and participating in this study. This work was entirely supported by the Center for Emerging Viral Diseases, a grant from the Private Foundation of the Geneva University Hospitals, the Pictet Charitable Foundation, the Ancrage Foundation and by research funds from the Center of Vaccinology.

## Bibliography

1. Amanat F, Krammer F. SARS-CoV-2 Vaccines: Status Report. Immunity. 2020 Apr 14;52(4):583–9.

2. Ohst C, Saschenbrecker S, Stiba K, Steinhagen K, Probst C, Radzimski C, et al. Reliable Serological Testing for the Diagnosis of Emerging Infectious Diseases. In: Hilgenfeld R, Vasudevan SG, editors. Dengue and Zika: Control and Antiviral Treatment Strategies [Internet]. Singapore: Springer Singapore; 2018 [cited 2020 Apr 22]. p. 19–43. (Advances in Experimental Medicine and Biology; vol. 1062). Available from: http://link.springer.com/10.1007/978-981-10-8727-1_3

3. Guidelines of the Office International des Epizooties for laboratory quality evaluation, for international reference standards for antibody assays and for laboratory proficiency testing. Rev - Off Int Epizoot. 1998 Aug;17(2):600–31.

4. Meyer B, Drosten C, Müller MA. Serological assays for emerging coronaviruses: Challenges and pitfalls. Virus Research. 2014 Dec 19;194:175–83.

5. Gorse GJ, Patel GB, Vitale JN, O’Connor TZ. Prevalence of antibodies to four human coronaviruses is lower in nasal secretions than in serum. Clin Vaccine Immunol. 2010 Dec;17(12):1875–80.

6. Müller MA, Meyer B, Corman VM, Al-Masri M, Turkestani A, Ritz D, et al. Presence of Middle East respiratory syndrome coronavirus antibodies in Saudi Arabia: a nationwide, cross-sectional, serological study. Lancet Infect Dis. 2015 Jun;15(6):629.

7. Corman VM, Müller MA, Costabel U, Timm J, Binger T, Meyer B, et al. Assays for laboratory confirmation of novel human coronavirus (hCoV-EMC) infections. Euro Surveill. 2012 Dec 6;17(49).

8. Okba NMA, Müller MA, Li W, Wang C, GeurtsvanKessel CH, Corman VM, et al. Severe Acute Respiratory Syndrome Coronavirus 2-Specific Antibody Responses in Coronavirus Disease 2019 Patients. Emerging Infect Dis. 2020 Apr 8;26(7).

9. Corman VM, Landt O, Kaiser M, Molenkamp R, Meijer A, Chu DK, et al. Detection of 2019 novel coronavirus (2019-nCoV) by real-time RT-PCR. Euro Surveill. 2020;25(3).

10. Sim J, Wright CC. The kappa statistic in reliability studies: use, interpretation, and sample size requirements. Phys Ther. 2005 Mar;85(3):257–68.

11. Berger Rentsch M, Zimmer G. A vesicular stomatitis virus replicon-based bioassay for the rapid and sensitive determination of multi-species type I interferon. PLoS ONE. 2011; 6(10):e25858.

12. Torriani G, Trofimenko E, Mayor J, Fedeli C, Moreno H, Michel S, et al. Identification of Clotrimazole Derivatives as Specific Inhibitors of Arenavirus Fusion. J Virol. 2019 15;93(6).

13. Fukushi S, Mizutani T, Saijo M, Matsuyama S, Miyajima N, Taguchi F, et al. Vesicular stomatitis virus pseudotyped with severe acute respiratory syndrome coronavirus spike protein. J Gen Virol. 2005 Aug;86(Pt 8):2269–74.

14. Glüer CC. Monitoring skeletal changes by radiological techniques. J Bone Miner Res. 1999 Nov;14(11):1952–62.

15. Seibel MJ, Koeller M, Van der Velden B, Diel I. Long-term variability of bone turnover markers in patients with non-metastatic breast cancer. Clin Lab. 2002;48(11–12):579–82.

16. DeLong ER, DeLong DM, Clarke-Pearson DL. Comparing the areas under two or more correlated receiver operating characteristic curves: a nonparametric approach. Biometrics. 1988 Sep;44(3):837–45.

17. Lassaunière R, Frische A, Harboe ZB, Nielsen AC, Fomsgaard A, Krogfelt KA, et al. Evaluation of nine commercial SARS-CoV-2 immunoassays. medRxiv. 2020 Apr 10;2020.04.09.20056325.

18. Duan J, Yan X, Guo X, Cao W, Han W, Qi C, et al. A human SARS-CoV neutralizing antibody against epitope on S2 protein. Biochem Biophys Res Commun. 2005 Jul 22;333(1):186–93.

19. Zhong X, Yang H, Guo Z-F, Sin W-YF, Chen W, Xu J, et al. B-cell responses in patients who have recovered from severe acute respiratory syndrome target a dominant site in the S2 domain of the surface spike glycoprotein. J Virol. 2005 Mar;79(6):3401–8.

20. Keng C-T, Zhang A, Shen S, Lip K-M, Fielding BC, Tan THP, et al. Amino Acids 1055 to 1192 in the S2 Region of Severe Acute Respiratory Syndrome Coronavirus S Protein Induce Neutralizing Antibodies: Implications for the Development of Vaccines and Antiviral Agents. J Virol. 2005 Mar;79(6):3289–96.

21. Zeng F, Hon CC, Yip CW, Law KM, Yeung YS, Chan KH, et al. Quantitative comparison of the efficiency of antibodies against S1 and S2 subunit of SARS coronavirus spike protein in virus neutralization and blocking of receptor binding: implications for the functional roles of S2 subunit. FEBS Lett. 2006 Oct 16;580(24):5612–20.

22. Omar F, Watt GF van der, Pillay TS. Reference change values: how useful are they? Journal of Clinical Pathology. 2008 Apr 1;61 (4):426–7.

